# Addressing the Slippery Slope Argument: Trends in Euthanasia Among Patients with Psychiatric Disorders and Dementia in Belgium, 2002–2023

**DOI:** 10.1101/2025.01.09.25320248

**Authors:** Jacques Wels, Natasia Hamarat

## Abstract

**Background:** Assisted suicide of patients with psychiatric disorders or dementia have increased in countries like Belgium, the Netherlands, and Canada and are often mentioned to argue against euthanasia regulations, suggesting they lead to a slippery slope. We examine changes in euthanasia cases for patients with these conditions in Belgium.

**Data and methos:** We use data on all cases of euthanasia reported to the Federal Commission for the Control and Evaluation of Euthanasia (FCCEE) from 2002 to 2023. Psychiatric disorders (N=427) and dementia (N=310) represent 1.27% and 0.92% of all cases, respectively. Using time-series Poisson regression, we model trends by first examining interactions between euthanasia reasons and year, then extending to three-way interactions with gender and region. We replicate the analyses with an offset to account for demographic changes.

**Results:** Euthanasia for psychiatric disorders and dementia showed distinct trends over time. Euthanasia for psychiatric disorders followed trends similar to the other types of euthanasia, while euthanasia for dementia showed a slight over-increase. Demographic change explains part of the increase in euthanasia for dementia and other reasons but not psychiatric disorders. While euthanasia rates for psychiatric disorders were initially higher for women, the trend for men is catching up over time. Regional trends indicate higher overall euthanasia rates in the Dutch-speaking population but with faster increases in the French-speaking population.

**Discussion:** The Belgian experience demonstrates that assisted dying laws can include non-terminal psychiatric conditions with appropriate safeguards and without evidence of significant misuse, addressing concerns about the slippery slope argument.

## Background

Research on assisted dying using actual data is sparse, [1] and much of the scientific literature— often consisting of opinion pieces [2,3] — relies on ethical debates, leaving little room for empirical evidence. Similarly, when data are mobilized, they are often examined at a highly descriptive level, focusing on assisted dying trends while overlooking changes in population structures during the selected period. The lack of rigor in addressing assisted dying trends in states that have implemented such legislation undermines the quality of the debate. This leaves room for fallacious arguments, such as the purported “slippery slope,” which is often mentioned but has never been substantiated [4–8].

The “slippery slope” argument is a common criticism of voluntary active euthanasia, suggesting that once euthanasia is permitted for specific conditions, it may lead to broader, less ethically acceptable practices, such as for non-terminal or psychiatric conditions [7,9]. This concern emphasises the need for rigorous safeguards, particularly in cases involving psychiatric disorders, to ensure mental competence in patients seeking euthanasia for physical ailments [10]. Additionally, critics argue that specific groups, including those in under-resourced healthcare settings [4] or deprived care facilities [9], could be disproportionately affected, raising concerns about potential coercion.

Previous research has particularly focused on gender and regional discrepancies. For instance, it was shown that female patients are notably overrepresented in psychiatric cases, although these comprise a small fraction of total cases [11,12]. Gender disparities have been noted in the context of Belgian euthanasia data, which shows a relatively balanced distribution with females representing 49.6 percent of euthanasia cases in 2020 [13] and data on euthanasia as the ratio on all deaths by gender show similar rates among genders [14]. Regional differences have also been documented. For example, in the Netherlands, unexplained geographical variations in euthanasia incidence were observed across provinces. Factors such as age, church attendance, political orientation, income, self-perceived health, and availability of voluntary workers have been associated with these differences, yet a significant portion of the variation remains unexplained [15]. In Belgium, official statistics reveal higher propensities for euthanasia in the Flemish region [16], with most research predominantly focused on Flanders [17,18].

Assisted dying, in this configuration, would not be efficiently monitored and controlled and might lead to error, abuse or violation of the rights of vulnerable patients [19]. While the slippery slope argument often focuses on potential abuses, evidence from countries where euthanasia is legalized indicates that although practices have expanded, they generally remain within legal and ethical boundaries [20]. Studies focusing on the slippery slope assumption rarely focus on data [6] and empirical investigation do not attest the existence of a slippery slope in the Netherlands [21] or Oregon [22].

Although there is no evidence on the existence of a slippery slope, the application of euthanasia for psychiatric disorders or dementia raises profound ethical concerns. Proponents argue that individuals suffering from severe mental illnesses, such as treatment-resistant depression or complex post-traumatic stress disorder, may experience unbearable suffering that justifies euthanasia [23]. However, critics contend that mental illness can impair judgment and decision-making, potentially compromising informed consent. The absence of universally accepted diagnostic criteria for the severity of psychiatric suffering further complicates these discussions, leading to calls for stringent safeguards to prevent abuse and ensure appropriate psychiatric care [24].

The ethical and legal frameworks governing euthanasia vary significantly across countries. In jurisdictions such as the Netherlands, Belgium, and Canada, euthanasia is legally permitted under specific circumstances. The Netherlands was the first country to legalize euthanasia in 2002, extending the law to include individuals with psychiatric conditions by 2014. Belgium, which followed, has allowed euthanasia since 2002 for both terminal and non-terminal conditions [25], including severe mental illness and dementia. Belgium’s inclusion of psychiatric disorders from the outset nuances the “slippery slope” argument, which suggests that permitting euthanasia for non-terminal illnesses inevitably follows the implementation of more restrictive regulations.

The Belgian Euthanasia Act allows legally competent adults experiencing “constant and unbearable physical or mental suffering that cannot be alleviated” due to a serious and incurable medical condition to request euthanasia. Safeguards exist. The request must be voluntary, deliberate, and repeated, and it must be made in writing. Regarding the expected time of death, one or two independent physicians − either psychiatrist or specialist of the pathology − who are not affiliated with the attending physician or the patient and are knowledgeable of the medical condition in question, must also be consulted.

The total number of reported cases of euthanasia was 235 in 2003 and 1,807 in 2013 [20]. They were 2,700 in 2021 [26]. Euthanasia is mostly linked to chronic or terminal physical conditions – with a large share due to cancers in terminal phase – but psychiatric disorders also lead to a substantial proportion of euthanasia [27]. It was estimated that, between 2002 and 2021, euthanasia for unbearable suffering caused by psychiatric disorders concerned 370 patients, 1.4 percent of the total number of euthanasia cases, most of which occurred after 2010. Research on medical files has found that most (90 percent) of these were diagnosed more than one disorder [28].

Using administrative data on all euthanasia cases reported to the Federal Control and Evaluation Commission on Euthanasia (FCCEE) over the past 22 years (2002–2023), this study aims to address the potential existence of a slippery slope in Belgium by answering two specific research questions: (R.Q.1) Are trends in euthanasia for psychiatric disorders and dementia more pronounced compared to those observed for euthanasia due to other reasons including terminal illness?; (R.Q.2.) Are specific trends evident among population sub-groups, such as gender and region?

## Data and methods

### Data

We use data routinely collected by the Federal Commission for the Control and Evaluation of Euthanasia (FCCEE), derived from individual reports submitted by euthanasia practitioners. These reports are fully anonymized and encompass all reported euthanasia cases since 2002, including information on the reasons for euthanasia, as well as the patients’ gender, age group, and language. Access to data was granted by the Federal Commission for the Control and Evaluation of Euthanasia (FCCEE) ethics committee on the 14^th^ of May 2024.

The dataset includes 33,647 cases, representing all reported euthanasia cases in Belgium between 2002 and 2023. Since the euthanasia law was implemented in mid-2002, we exclude data from 2002 in our empirical models because the law was implemented in mid-2002 leading to low cases (N=24). Additionally, 43 cases were removed due to incomplete information. No imputations were made to address the missing data, given the small proportion (0.1% of the total) and limited available information. The final sample includes 33,623 cases. Among these, 427 cases were justified by psychiatric disorders and 310 cases by dementia, i.e., respectively 1.27 and 0.92 percent of all cases of euthanasia observed over the period.

The dataset contains aggregated count data stratified by multiple demographic, temporal, and contextual factors. Each row represents a unique combination of attributes, including year, language, gender, age, age group, location, and reason for the case. The primary variable of interest is the count of cases for each combination of factors, with an additional variable providing the total count of cases aggregated by year. The dataset is structured as a rectangular table, with each row corresponding to a unique combination of predictor variables, ensuring that all possible combinations are represented, including those with zero counts.

### Reasons for euthanasia

The Federal Commission for the Control and Evaluation of Euthanasia (FCCEE) identifies twelve medical conditions that may justify euthanasia (post-coded based on specific conditions mentioned by the medical practitioner on the euthanasia certificate). In this study, we classify these reasons into seven categories: (1) cancer and tumours; (2) multimorbidity; (3) nervous system diseases; (4) specific diseases; (5) psychiatric disorders; (6) cognitive disorders (e.g., dementia); and (7) others. The “specific diseases” category encompasses conditions of the respiratory, circulatory, genitourinary, and digestive systems, as well as haematological, endocrine, nutritional, and metabolic disorders. It also includes diseases of the eye, ear, musculoskeletal system, skin, and subcutaneous tissue. The “others” category includes abnormal clinical and laboratory findings not elsewhere classified, traumatic injuries, poisonings, consequences of external causes, perinatal conditions, congenital malformations, chromosomal abnormalities, and certain infectious or parasitic diseases.

We maintain the FCCEE’s distinction between psychiatric and cognitive disorders due to their differing clinical profiles. For analytical purposes, we group the reasons for euthanasia into three broad categories: (1) psychiatric disorders, (2) dementia, and (3) all other justifications, excluding psychiatric disorders and dementia.

### Covariates

The dataset includes several key variables for analysis. The year of euthanasia, recorded from 2003 to 2023 (we removed the year of the implementation of the law as it does not cover the full 2002 year), is used as a continuous variable (coded 0–21) in main analyses, with a sensitivity check treating it as categorical to address non-linear trends. Age is categorized into eight groups—15–29, 30–39, 40–49, 50–59 (reference), 60–69, 70–79, 80–89, and 90+—to ensure anonymity. Gender, reported by the medical practitioner, is recorded as male or female (reference). To account for regional differences within Belgium’s federal structure, we include language data (Dutch or French, reference) used by the reporting practitioner, as place of residence was inconsistently collected. Additional variables include the basis for euthanasia (advanced request or actual, reference), type of unbearable suffering (physical, reference; mental; or both), term of death (expected within a year or longer, reference), and place of death, categorized as home (reference), hospital, care home, palliative care, or other. These variables are systematically included.

### Population offset

We generate population figures based on demographic data retrieved from *Statbel*. This data includes information on the total population as of January 1st for each selected year (2002 to 2023), broken down by age group, sex, and region of residence. We chose to use population figures instead of the number of deaths by sub-group, as done in previous studies [8,15,29,30], because a non-negligible share of euthanasia is performed on patients not expected to die in the foreseeable future, including those with dementia or psychiatric disorders – 14.4 percent of all cases in 2020-2021 [13]. The figures are calculated for each line of euthanasia counts by year, age, gender, and language, and are then used as offset in the model. Demographic data do not include information on language. To tackle this issue, the French-speaking population was calculated as the sum of the population residing in Wallonia and 90 percent of the population in Brussels and the Dutch speaking as the sum of the Flanders residents and 10 percent of the Brussels population, reflecting the Belgian language repartition.

### Analyses

We use a Poisson fixed effects analysis on the count data of euthanasia cases in Belgium, examining the relationship of year, age group, gender, language and reason with euthanasia counts. Since the study examines count data we apply a Poisson model, which is specifically designed for non-negative integer outcomes and accounts for the mean-variance relationship inherent in such data [31], as done previously on suicide count data [32,33]. We compare two models. The first model does not include an offset for population size, and thus estimates the *occurrence* of euthanasia, providing insights into the raw counts without accounting for differences in population size. The second model includes an offset for population size by year, age, gender, and region, allowing us to calculate the *prevalence* of euthanasia, i.e., the rate of euthanasia occurrences relative to the population at risk. We exponentiate the coefficients to obtain the rate ratios (RR) and prevalence ratios (IR) [34,35]. The analysis is not based on a population sample, and significance levels are not required. We replicate the analyses with (a) a two-way multiplicative interaction between year and reason controlling for gender and region; (b) a three-way interaction between year, reason and gender controlling for region; (c) and a three-way interaction between year, reason and region controlling for gender. All analyses reported in this study were made using the R software, using especially the “glm” and “predict” packages.

## Results

Table 1 exhibits the number of euthanasia cases by reason, distinguishing between dementia, psychiatric disorders, and other causes from 2002 to 2023. To ensure anonymity and avoid low counts (<3), the years 2002 to 2006 (the first five years following legalization) are grouped into a single category. Euthanasia cases for dementia or psychiatric disorders are reported 100 times less frequently than for other reasons, such as cancer or comorbidities, while following a similar trend to these more common cases.

**Table 1.** Yearly count of reported cases of euthanasia cases by reason.

| Year | Terminal illness |  | Dementia |  | Psychiatric disorders |  | Total |
| --- | --- | --- | --- | --- | --- | --- | --- |
|  | Count | Percent | Count | Percent | Count | Percent |  |
| 2002-2006 | 1,417 | 99.23 | 5 | 0.35 | 6 | 0.42 | 1,428 |
| 2007 | 491 | 99.80 | 1 | 0.20 | 0 | 0.00 | 492 |
| 2008 | 695 | 98.86 | 5 | 0.71 | 3 | 0.43 | 703 |
| 2009 | 804 | 98.17 | 6 | 0.73 | 9 | 1.10 | 819 |
| 2010 | 928 | 98.10 | 8 | 0.85 | 10 | 1.06 | 946 |
| 2011 | 1,099 | 97.52 | 14 | 1.24 | 14 | 1.24 | 1,127 |
| 2012 | 1,384 | 96.92 | 14 | 0.98 | 30 | 2.10 | 1,428 |
| 2013 | 1,760 | 97.24 | 13 | 0.72 | 37 | 2.04 | 1,810 |
| 2014 | 1,866 | 96.78 | 18 | 0.93 | 44 | 2.28 | 1,928 |
| 2015 | 1,959 | 96.88 | 20 | 0.99 | 43 | 2.13 | 2,022 |
| 2016 | 1,988 | 98.03 | 11 | 0.54 | 29 | 1.43 | 2,028 |
| 2017 | 2,273 | 98.27 | 14 | 0.61 | 26 | 1.12 | 2,313 |
| 2018 | 2,303 | 97.63 | 22 | 0.93 | 34 | 1.44 | 2,359 |
| 2019 | 2,608 | 98.16 | 26 | 0.98 | 23 | 0.87 | 2,657 |
| 2020 | 2,401 | 98.20 | 23 | 0.94 | 21 | 0.86 | 2,445 |
| 2021 | 2,647 | 98.11 | 27 | 1.00 | 24 | 0.89 | 2,698 |
| 2022 | 2,898 | 97.71 | 42 | 1.42 | 26 | 0.88 | 2,966 |
| 2023 | 3,334 | 97.40 | 41 | 1.20 | 48 | 1.40 | 3,423 |

### R.Q.1 Are trends in euthanasia for psychiatric disorders and dementia more pronounced compared to those observed for euthanasia due to other reasons?

We present the exponentialized coefficients from Poisson regression with (RR, Relative Risk) and without (IR, Incidence Rate) a demographic offset in table 2. The main model incudes a two-way multiplicate interaction term between the year cases were reported (from 0 to 21) as numeric and reason for euthanasia. Other models use a three-way interaction including, respectively, gender, region, basis, expected term of death, type of suffering and place of death. Table 2 exhibits both the main effects and the interaction terms whilst figure 1 shows the predicted IR and RR for each model (except the type of suffering and the place of death that include too many modalities). Full results presenting the exponentiated estimates are shown in <u>supplementary file S1</u>.

**Figure 1.**
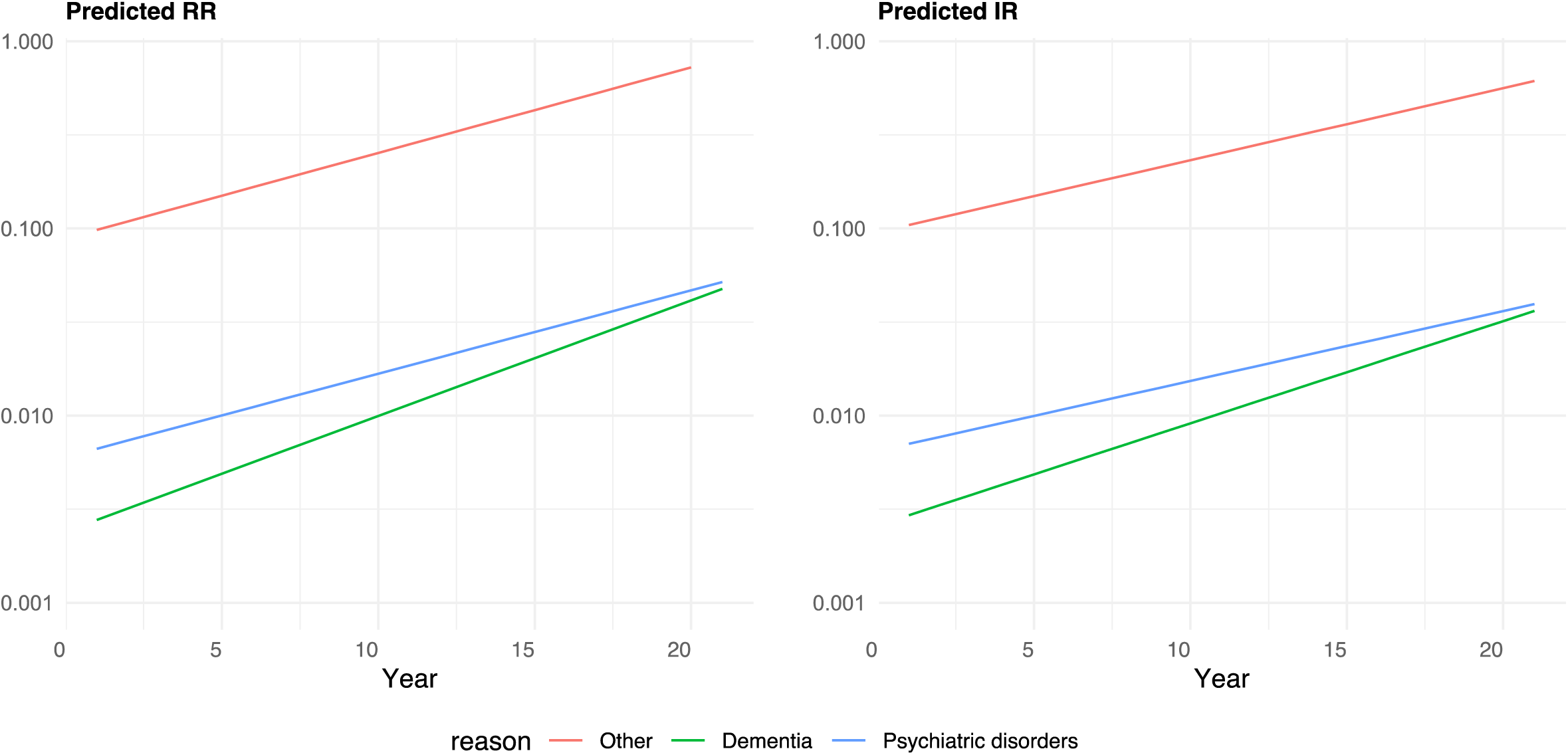
Predicted Relative Risks and Incidence Ratios of euthanasia for dementia, psychiatric disorders and other causes, 2003 to 2023.

**Table 2.**
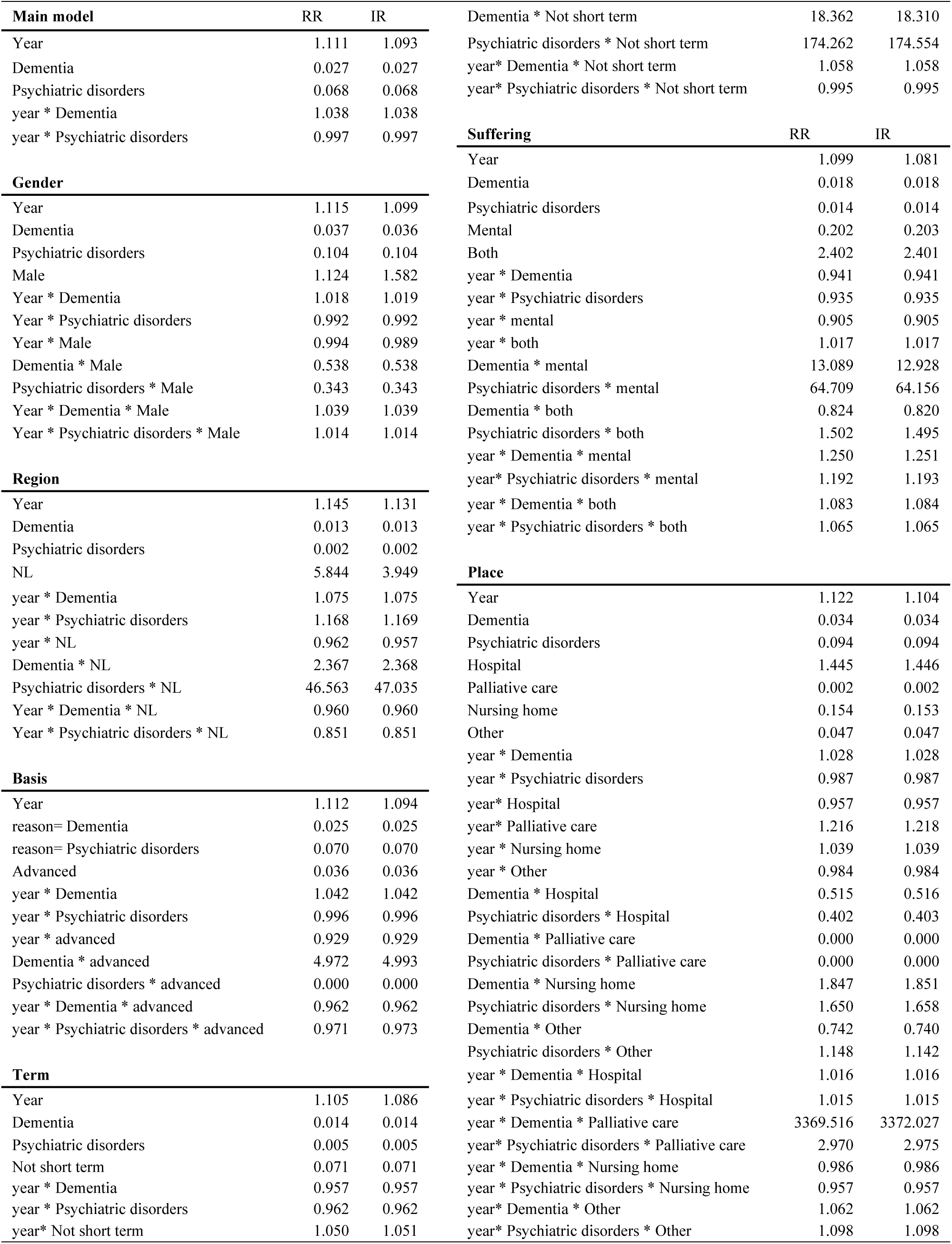
Relative risks (RR) and Incidence Ratios (IR) of the two-way and three-way interaction terms.

The main model investigated the two-way interaction between the reason for euthanasia and time, comparing trends for psychiatric disorders and dementia to “other” reasons, which serve as the reference category. The findings indicate that euthanasia for dementia has shown a modest but significant annual increase relative to “other” reasons (year × dementia, RR = 1.038; IR = 1.038). This result suggests that the practice of euthanasia for dementia-related cases is becoming relatively more common over time compared to “other” reasons. In contrast, the trend for psychiatric disorders remains stable, as evidenced by the interaction between time and psychiatric disorders (year × psychiatric disorders, RR = 0.997; IR = 0.997). This finding highlights that psychiatric disorders have not followed the same increasing trajectory as dementia when compared to “other” reasons. We also observe, as can be seen in figure 1, that not accounting for population composition and change leads to overestimate euthanasia prevalence in Belgium. However, including a demographic adjustment does not change the trends observed for the different types of euthanasia.

### R.Q.2. Are specific trends evident among population sub-groups, such as gender and region?

The gender-specific model revealed notable differences in trends. For dementia, the temporal increase compared to “other” reasons was slightly more pronounced among males (year × dementia × male, RR = 1.039). However, psychiatric disorders in males exhibited a lower relative risk compared to “other” reasons (year × psychiatric disorders × male, RR = 0.343), suggesting that the risk and prevalence of euthanasia for psychiatric disorders relative to “other” reasons are reduced in males. T

Regional patterns between the Dutch-speaking northern region (NL) and the French-speaking southern region (FR) also emerged. Dementia-related euthanasia in the NL region showed a significant relative increase compared to “other” reasons (year × dementia × NL, RR = 2.367). Conversely, psychiatric disorders in the NL region exhibited a relative stable trend with a slope lower compared to the “other” reasons (year × psychiatric disorders × NL, RR = 0.851). Euthanasia case are less prevalent in the FR region but the slopes observed for each types indicate a greater increase in the south of the country.

The analysis of the basis of euthanasia revealed distinct trends. For planned euthanasia, dementia demonstrated higher prevalence compared to the “other” reasons (year × dementia × planned, RR = 4.972). In contrast, psychiatric disorders exhibited no significant relative increase for planned euthanasia compared to “other” reasons (year × psychiatric disorders × planned, RR = 0.971).

Dementia cases involving mental suffering showed significantly higher relative risks compared to “other” reasons (dementia × mental suffering, RR = 13.089), suggesting that mental suffering substantially impacts the likelihood of euthanasia in dementia cases. Similarly, psychiatric disorders involving both physical and mental suffering demonstrated high relative risks (psychiatric disorders × both types of suffering, RR = 64.709), reflecting the multifaceted considerations in these cases.

The three-way interaction with the place of death is less easy to interpret given the low count observed of euthanasia for psychiatric disorders and dementia. What can be observed is that euthanasia for both reasons is more prevalent at home than in hospital or palliative care settings. However, there has been an increase over time of euthanasia for dementia and psychiatric disorders in palliative care (low count must prevent to call it a trend). Unsurprisingly, euthanasia for dementia is more likely to occur in a nursing home than at home (RR = 1.847) but this is also the case of euthanasia for psychiatric disorders (RR = 1.650). These findings remained stable over the years.

**Figure 2.**
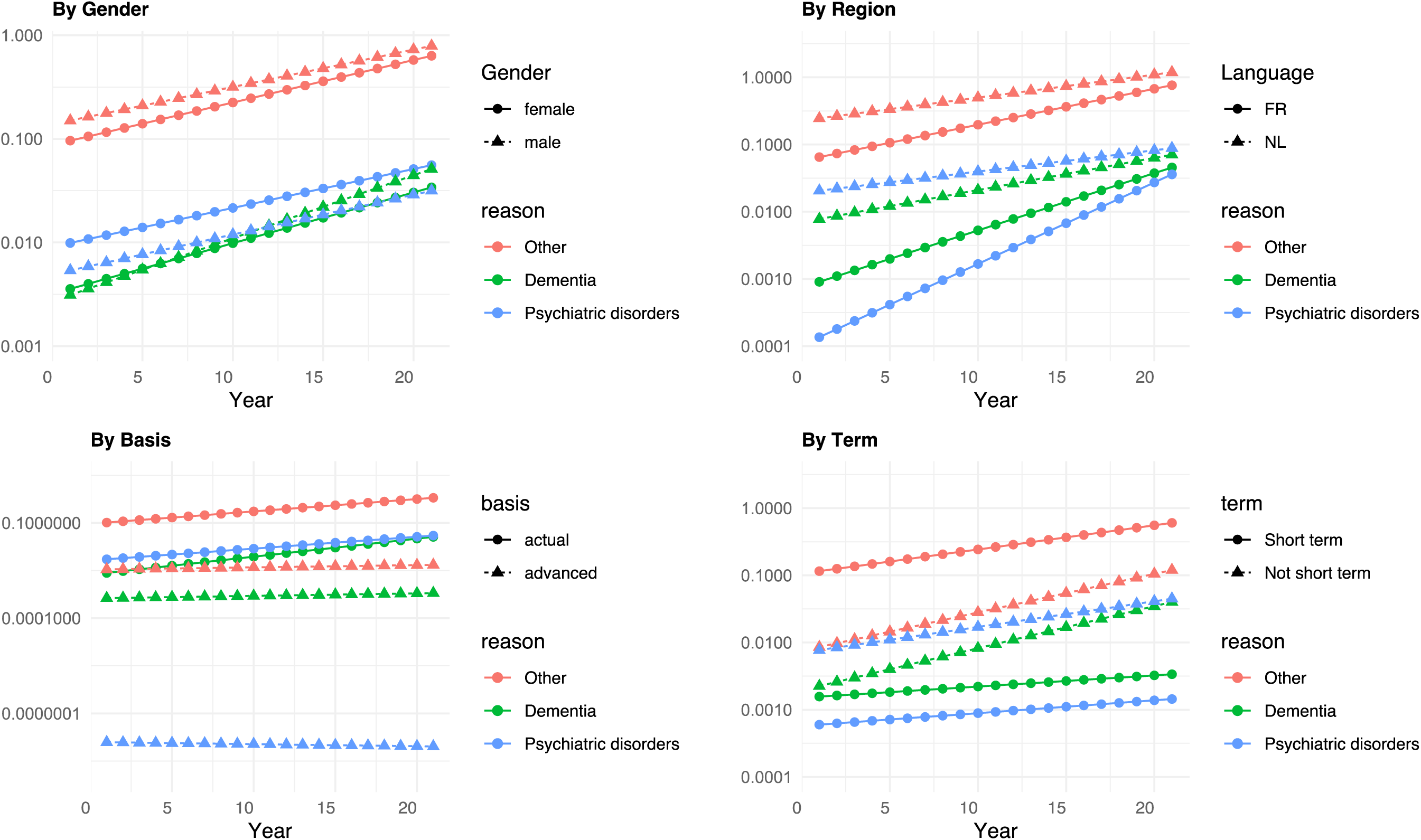
Predicted Incidence Ratios of euthanasia by reason, year and gender, region, basis and term, 2003 to 2023.

The temporal trends indicate that dementia-related euthanasia consistently increased relative to “other” reasons across all models, particularly in nursing homes, for planned cases, and for cases involving mental suffering. By contrast, psychiatric disorders remained stable, with no significant increase compared to “other” reasons across the models. These findings suggest that dementia-related euthanasia is becoming relatively more common, whereas euthanasia for psychiatric disorders shows no evidence of relative growth over time. To confirm these trends, we have addressed non-linearity in euthanasia trends using year as a factor variable with no major differences in findings, as can be seen in <u>supplementary files 4-6</u>.

## Discussion

Recent debates on the implementation of assisted dying in the United Kingdom and France have highlighted the importance of restricting the right to euthanasia to patients with terminal physical illnesses, excluding psychiatric disorders or degenerative conditions such as dementia.

Opponents of such rights often argue that a narrowly defined assisted dying law would inevitably expand to include non-terminal illnesses, resulting in an uncontrolled rise in cases. The Belgian experience, however, addresses these concerns. From the outset, the Belgian law has allowed euthanasia for non-terminal and non-physical illnesses under strict safeguards, enforced by medical practitioners and monitored by the Federal Commission for the Control and Evaluation of Euthanasia.

Empirical evidence demonstrates no dramatic increase in euthanasia for non-terminal conditions between 2003 and 2023: psychiatric disorders and dementia accounted for only 1.4% and 1.2% of all cases in 2023, respectively. Trends show gradual implementation, with a steady yearly rate of change for psychiatric conditions and a slightly sharper but still modest increase for dementia. During the first five years following the implementation of the regulation, cases of euthanasia for psychiatric disorders and dementia remained rare, indicating that medical practitioners were reluctant at first to practice euthanasia on patients suffering from cognitive or psychiatric conditions. Differences in access across genders and linguistic regions have narrowed over time, suggesting harmonization rather than segmentation.

Request for euthanasia made in advance are rare and mainly associated with non-psychiatric and non-dementia conditions. Euthanasia on patients not expected to die in the foreseeable future have risen more sharply among those with ‘other” conditions groups rather than among those with psychiatric conditions. However, we observe a sharp increase in euthanasia for patients with dementia not expected to die in the forceable future. As cases remain low, it is difficult to draw any conclusion on future trends but euthanasia seems to become more common among patients with dementia, living in care how and not expected within a year.

Nonetheless, Belgium faces significant challenges in evaluating equity in access to euthanasia. The absence of social security identifiers in euthanasia records prevents data linkage with socio-economic information, while a lack of geographic data precludes sub-regional comparisons. These gaps are compounded by the limited annual budget of €160,000 allocated to the Federal Commission, constraining its ability to support robust research and comprehensive evaluation. Addressing these issues requires proactive planning, including establishing data collection protocols, securing patient consent, ensuring data anonymization, enabling data linkage, and committing to independent research. Such measures are essential for monitoring trends, ensuring equitable access, and preventing potential discrimination.

Nevertheless, the Belgian case provides compelling evidence against the slippery slope argument, which has often been invoked to oppose legislation expanding individual rights, such as those related to reproductive health, same-sex marriage, or abortion. While theoretical concerns justify the inclusion of safeguards, empirical evidence does not support fears of unchecked expansion or abuse in euthanasia cases. Instead, the slippery slope argument should be regarded as a speculative myth rather than a substantiated fact. Policymakers should prioritize designing thoughtful, well-regulated frameworks informed by evidence as well as proper monitoring strategies rather than being deterred by unfounded concerns.

## Data Availability

Access is be granted upon request to the Federal Commission for the Control and Evaluation of Euthanasia (FCCEE): https://consultativebodies.health.belgium.be/en/advisory-and-consultative-bodies/federal-commission-control-and-evaluation-euthanasia

